# Children’s visual acuity tests at home: A prospective repeated measures study

**DOI:** 10.1101/2022.10.14.22281044

**Authors:** Daniel Osborne, Aimee Steele, Megan Evans, Helen Ellis, Roshni Pancholi, Tomos Harding, Jessica Dee, Rachel Leary, Jeremy Bradshaw, Elizabeth O’Flynn, Jay E Self

**Affiliations:** University Hospital Southampton NHS Foundation Trust, Department of Ophthalmology, Southampton, UK; University of Southampton Faculty of Medicine, Southampton, UK

**Keywords:** Children, Amblyopia, Paediatric ophthalmology

## Abstract

**Background:** Home visual acuity tests could ease pressure on ophthalmic services by facilitating remote review of a variety of patients. Home tests may have further utility in giving service users frequent updates of vision outcomes during therapy, identifying vision problems in an asymptomatic population, and engaging stakeholders in therapy.

The accuracy of home vision tests for children when completed without supervision from a professional is unknown.

**Methods:** Children attending outpatient clinics had their visual acuity measured 3 times in a randomised order at the same appointment. Once by a registered orthoptist as per standard clinical protocols, once by an orthoptist using a tablet-based visual acuity test (iSight Pro, Kay Pictures), and once by an unsupervised parent or carer using the tablet-based test.

**Results:** 42 children were recruited to the study. The mean age was 5.6 years (range 3.3 to 9.3 years). Median measurements (interquartile range) for clinical standard, orthoptic-led and parent/carer-led iSight visual acuity measurements were 0.155 (0.18), 0.180 (0.26), and 0.300 (0.33) respectively.

The iSight app in the hands of parents/carers was significantly different from the standard of care measurements (P=0.009). In the hands of orthoptists, there was no significant difference between the iSight app and standard of care (P=0.551), nor was there significant difference between parents/carers using the app and orthoptists using the app (P=0.133).

**Conclusion:** This technique of unsupervised home visual acuity measures for children is not comparable to clinical measures and is unlikely to be valuable to clinical decision making and screening. Future work should focus on improving the technique through, for example, gamification of vision tests.

**What is already known on this topic?:** Children’s game visual acuity tests could improve things for hospital services and patients. The tablet-based tests are accurate when used by professionals to tests adults’ vision.

**What this study adds:** This study provides data about the accuracy of home tests when used by untrained parents or carers on their children.

**How this study might affect research, practice, or policy:** Current policy recommends apps home tests are not used to test children’s eyes for clinical decision-making purposes. Our data supports this policy and highlights the need for future research to focus on improving the tests.

**Synopsis / precis (“At a glance”):** Visual acuity testing at home could improve patient care and reduce clinical visits, but data to show clinicians that they can rely upon the results in children are lacking. Our study shows the tests cannot be relied upon for clinical purposes. Improvements to the tests are required before they become useful and can be implemented into practice.

## Introduction

Visual acuity is a clinical measure of how small a black-on-white object an eye can resolve. It is commonly used to screen for eye conditions, monitor disease progress, classify disease severity, and tailor treatment plans. Children’s visual acuity requires specialist equipment and a trained professional. Inaccurate measurement may result in false negatives/positives in screening programmes, misdiagnosis, or over or under treatment.

Up to 5% of children have amblyopia, which requires 6 to 8-weekly visits to a centre for a visual acuity test (1,2). Children that require repeat hospital visits have poor attendance at school, which may impact on their social and emotional development. Furthermore, repeat visits to the hospital places financial and time strain on parents or carers.

The UK National Screening Committee (NSC) recommends school-entry (age 4 to 5 years) vision screening, requiring every child to achieve a visual acuity of 0.20 logMAR or better to pass (3). Children with poorer visual acuity are referred to hospital services. With school closures during 2020-21 academic year, vision screening has had a reduced capacity.

The National Health Service (NHS) spends approximately £1365 on treating each child with amblyopia (2). Furthermore, most NHS Trusts have backlogs of amblyopia patients awaiting treatment, having cancelled outpatient follow-up for patients during lockdowns in 2020 (4).

Clinicians and professionals have long suggested home visual acuity tests may help to address these problems experienced by patients and NHS services. However, home testing has not yet made it to widespread use. There is concern that the tests may not be accurate enough to be relied upon when considering whether to see a patient in clinic, or to start or end therapy (5). Inaccuracies could be caused by change in test distance, peeking around occlusive glasses / eye patch, examiners offering children cues, or early termination of the test as the child loses interest.

Computerised visual acuity tests (apps) have previously been compared to their traditional printed equivalents (6). This study showed that the apps were comparable to validated printed tests. However, both VA tests in this study were led by a trained orthoptist rather than by parents / carers, as they would be at home. A letter based, home vision test has been developed and tested in a home setting on children (7,8). The system uses two computer devices to ensure correct test distance is maintained throughout examination, which is completed by parents on their children with supervision from a professional via video link. There was agreeability between video-supervised, parent self-test of their child and clinical measurements.

The primary aim of our study was to collect data about the accuracy of parent / carer-led visual acuity tests of their children using widely available tablet-based software (apps). A secondary outcome was to collect quantitative data about families’ access to the required equipment and technology. Additionally, in a questionnaire, we asked families if they found the tests easy and whether the outputs of the data would be helpful to them during their child’s care.

## Materials and Methods

### Study design and participants

Children between age 3 and 10 years were identified from orthoptic and paediatric ophthalmology outpatient clinics by a member of the research team (DO, ME, or AS). Recruitment ran from July 2020 to November 2021 and a sampling was dependant on availability of research staff, equipment, and outpatient clinic capacity. Children were excluded if they were unable to complete a clinical subjective visual acuity test, if their parent or carer was under age 17 years (a regulatory requirement of the Kay Pictures iSight Pro software), or they or their parent / carer was not willing to give informed consent.

Upon consent, children were assigned a sequential, unique study identifier (USI) number. Each participant completed three visual acuity tests on the same day in the clinic office:

a. A standard of care, clinical visual acuity test. A registered orthoptist tested visual acuity as per clinical guidelines and experience. Orthoptists selected an age and level of understanding appropriate test from: Single of Linear Crowded Kay Pictures (Tring, UK) book, Keeler (Windsor, UK) logMAR book or Bailey-Lovie letters on a Thomson (Welham Green, UK) Test Chart.
b. An orthoptist-led, tablet computer-based (Apple iPad, Apple, California, USA; iSight Pro, Kay Pictures, Tring, UK), visual acuity test. The same orthoptist that completed the standard of care test measured the participant’s visual acuity using the iSight app and an iPad. As they completed the test (approximately 10 minutes), they showed the parent or carer how to:
  – load the application
  – select the appropriate visual acuity test
  – Measure the correct distance to perform the test
  – Effectively occlude one eye (using either occlusive glasses or Durapore over one lens of spectacles) for uniocular testing
  – Complete the test and record the result
c. A parent led, tablet computer-based visual acuity test. Following observation of use of the iPad, the parent was asked to measure their child’s visual acuity using the iSight app. They were left alone in a clinical room for up to 15 minutes. The room had a 3-metre distance from the patient marked on the floor.

Children with odd USI numbers completed the tests in the order they appear above (*abc*), whereas those with even USI numbers used a *bac* order. This aimed to reduce order effects as children tire through the testing procedures but give parents/carers opportunity to see the iSight Pro app in use prior to using it themselves. Following completion, the participant returned to clinical care, the parent or carer completed a short questionnaire (Supplementary Material 1), and visual acuity results were collated for analysis.

### Ethics

The study adhered to the tenets of the Declaration of Helsinki. The study was approved by the NHS Health Research Authority (HRA) and an NHS Research Ethics Committee (Queen Square NHS REC, Reference: 20/HRA/2585). Parents or carers gave written informed consent for their child to take part in the study.

### Statistical analysis

Comparisons between standard of care, orthoptist-led iSight, and parent / carer iSight tests were made using analysis of variance (ANOVA). Additionally, we used tests for correlation to assess the effect of age on test accuracy. The parent / carer’s iSight test’s application as a screening tool was assessed using a receiver operating characteristic (ROC) curve. We present the sensitivity-specificity of the parent / carer’s test in detecting true (as measured by clinical standard tests) reduced (greater than 0.20 logMAR) visual acuity.

## Results

### Characteristics of participants

42 children were recruited to the study (*Table 1*). The mean age was 5 years 7 months (SD 15 months, range 3 years 4 months to 9 years 4 months). 25 (59.5%) were male, 13 were under frequent outpatient follow-up for amblyopia therapy (occlusion or atropine penalisation therapy). Visual acuity data were collected for both eyes for all participants in accordance with the testing protocol.

**Table 1.** Demographics and characteristics of participants.

|  |  |  |
| --- | --- | --- |
| **Sex ( M / F )** |  | 25 / 17 |
| **Age at testing**<br>(Mean ± SD, [range]) | | 5 years 7 months ± 15 months,<br>[41 – 113 months] |
| **Ocular diagnosis**<br>N (%) | Amblyopia | 13 (31) |
|  | Hypermetropia | 13 (31) |
|  | No ocular disease | 4 (9.5) |
|  | Astigmatism | 3 (7.1) |
|  | Nystagmus | 3 (7.1) |
|  | Myopia | 2 (4.8) |
|  | Intermittent distance<br>exotropia | 2 (4.8) |
|  | Infantile cataract | 1 (2.4) |
|  | Vernal keratoconjunctivitis<br>(VKC) | 1 (2.4) |

### Differences between tests

The median values [25th-75th percentile] for the clinical standard, orthoptist iSight and parent iSight were 0.155 [0.095-0.275], 0.180 [0.100-0.360], and 0.300 [0.135-0.465] respectively (Table 2, Figure 1). The iSight app in the hands of parents/carers was significantly different from the standard of care measurements (P=0.009). In the hands of orthoptists, there was no significant difference between the iSight app and standard of care (P=0.551), nor was there significant difference between parents/carers using the app and orthoptists using the app (P=0.133). Bland-Altman plots show greater variation of the differences between parent/carer iSight and standard care than between orthoptist iSight and standard care (Figure 2).

**Table 2.**
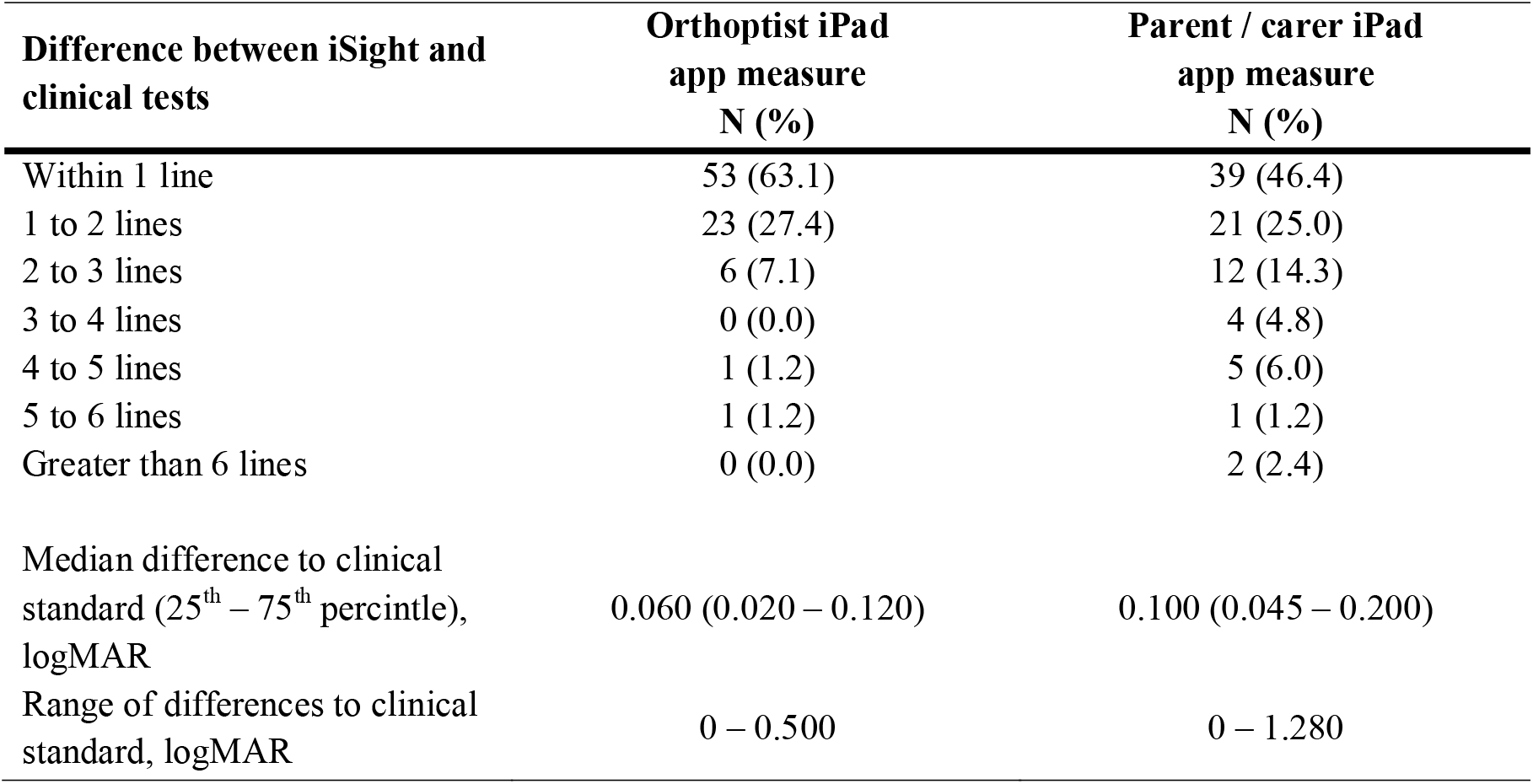
Orthoptists and parent / carer’s iSight measurements compared to standard clinical care measurements. N = 84 eyes, 42 participants.

| **Difference between iSight and**<br>**clinical tests** | **Orthoptist iPad**<br>**app measure**<br>**N (%)** | **Parent / carer iPad**<br>**app measure**<br>**N (%)** |
| --- | --- | --- |
| Within 1 line | 53 (63.1) | 39 (46.4) |
| 1 to 2 lines | 23 (27.4) | 21 (25.0) |
| 2 to 3 lines | 6 (7.1) | 12 (14.3) |
| 3 to 4 lines | 0 (0.0) | 4 (4.8) |
| 4 to 5 lines | 1 (1.2) | 5 (6.0) |
| 5 to 6 lines | 1 (1.2) | 1 (1.2) |
| Greater than 6 lines | 0 (0.0) | 2 (2.4) |
| Median difference to clinical<br>standard (25<sup>th</sup> – 75<sup>th</sup> percintle),<br>logMAR | 0.060 (0.020 – 0.120) | 0.100 (0.045 – 0.200) |
| Range of differences to clinical<br>standard, logMAR | 0 – 0.500 | 0 – 1.280 |

**Figure 1.**
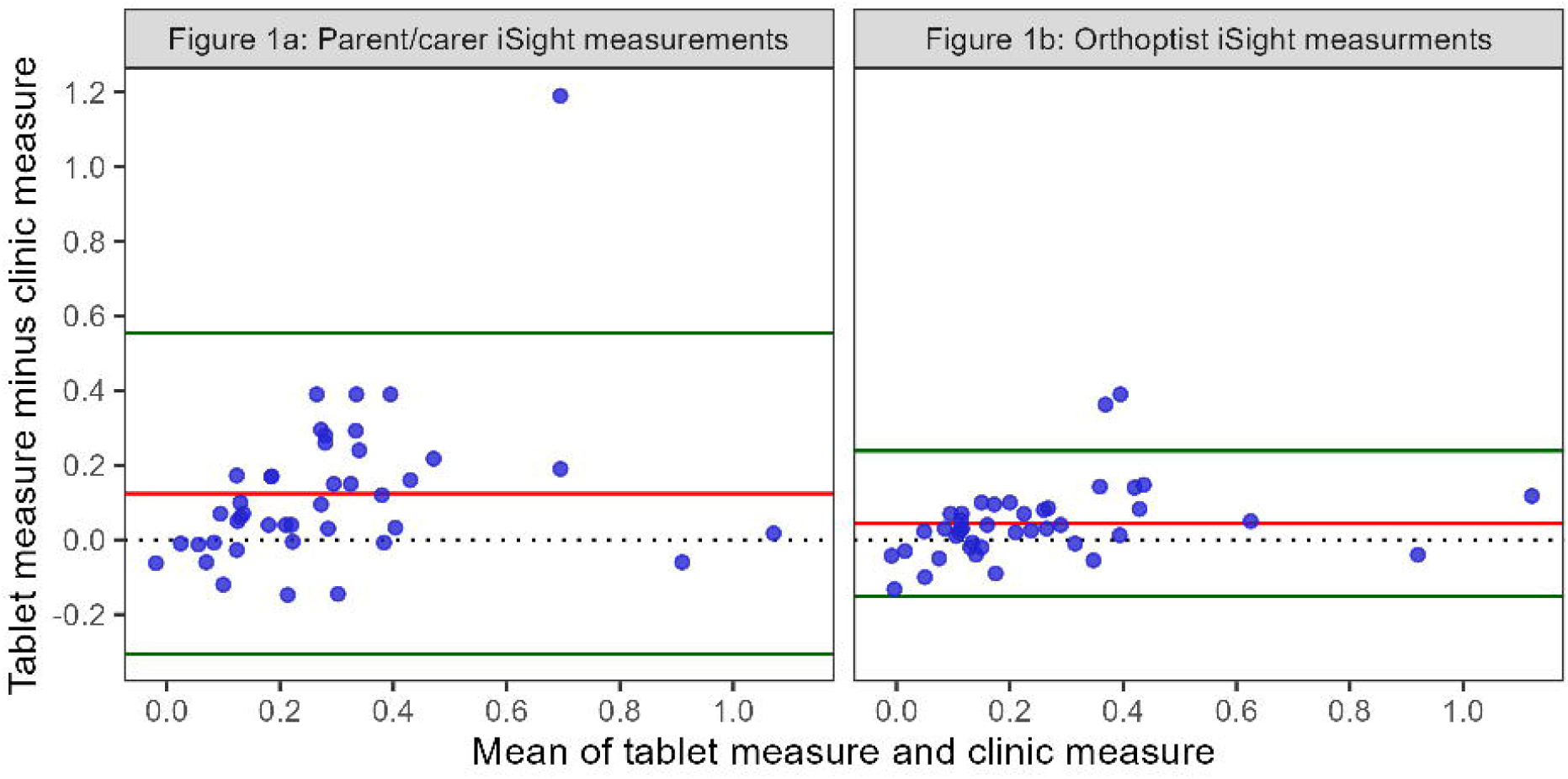
Boxplot showing visual acuity measures by testing method

**Figure 2.**
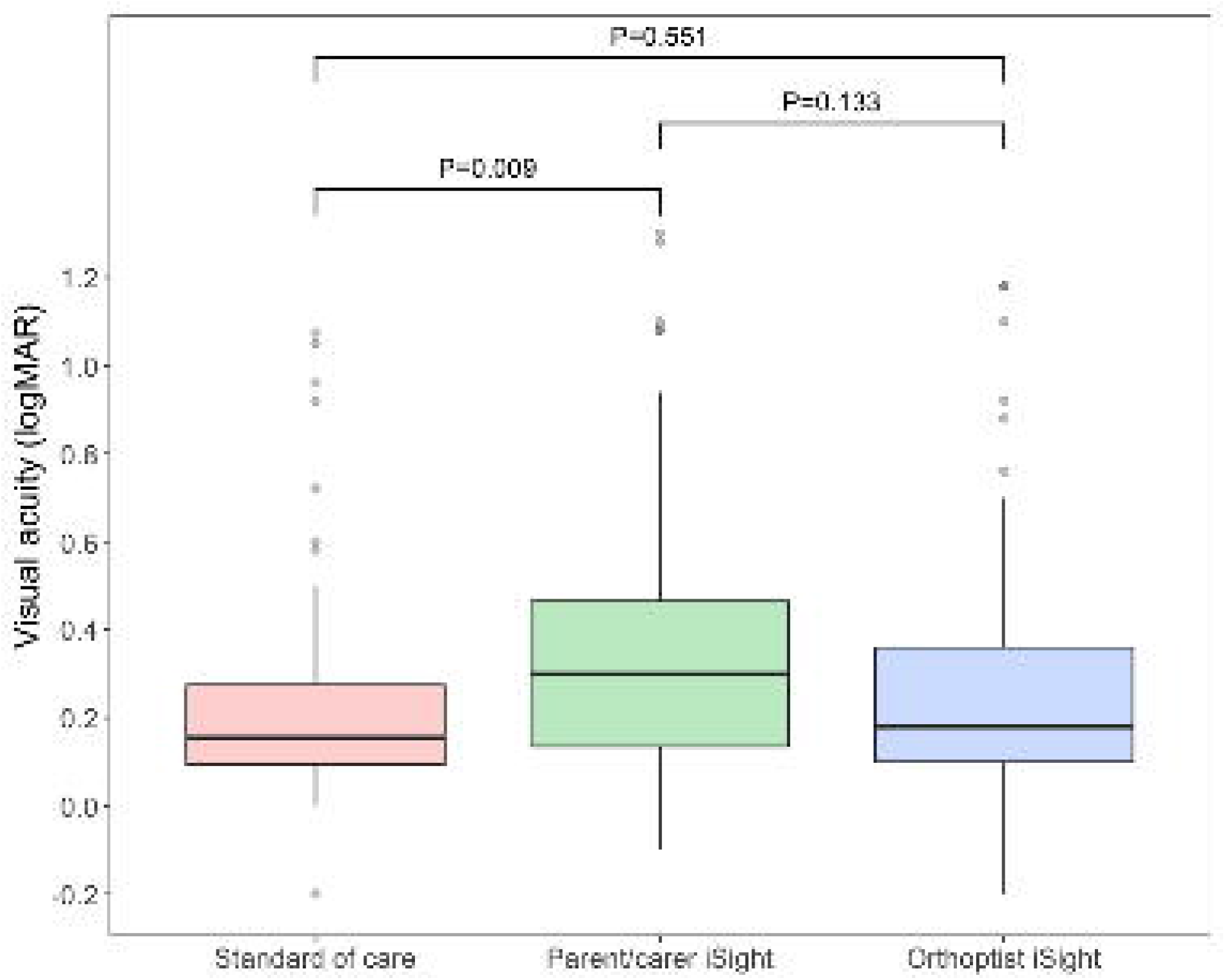
Bland-Altman plots comparing tablet-measured visual acuity to standard of care visual acuity. Red line denotes mean difference (bias), green lines denote mean difference ± 1.96 standard deviations (limit of agreement).

### Correlation between level of visual acuity and accuracy of parent / carer test

There was no correlation between worsening visual acuity and difference between clinic test and parent/carer iSight test (r=0.079, P=0.473).

### Parent / carer test as a screening tool

15 of 16 children with visual acuity equal to or worse (greater than) 0.20 logMAR in their worse seeing eye, as measured with standard clinical tests, did not achieve better than (less than) 0.20 logMAR on the parent / carer test (sensitivity = 93.8%). 12 of the 26 children with true visual acuity in their worse seeing eye better than or equal to 0.20 logMAR (measured with standard clinical tests), did not achieve 0.20 logMAR or better with the parent / carer test (false positive rate = 46.2%; specificity = 53.8%). Table 3 shows the sensitivity-specificity confusion matrix. The area under the receiver operating characteristic (ROC) curve (supplementary material 2) was 0.812 (95% confidence interval = 0.722-0.903).

**Table 3.** Sensitivity and specificity of parent / carer visual acuity (VA) test compared to standard of clinical care. (pos = positive, neg = negative)

|  |  | True result (measured by standard clinical care tests) |  |  |
| --- | --- | --- | --- | --- |
| Parent / carer test result | *N = 42 participants* | Positive (VA > 0.20) | Negative (VA ≤ 0.20) | *Total* |
|  | Positive (VA > 0.20) | **15**<br>True pos | **12**<br>False pos | *27* |
|  | Negative (VA ≤ 0.20) | **1**<br>False neg | **14**<br>True neg | *15* |
|  | *Total* | *16* | *26* |  |

Participant 41 was the only false negative in our analysis. The clinic visual acuity was 0.200 and 0.375 in the right and left eyes respectively. The orthoptist iSight measured 0.34 and 0.52, and the parent / carer iSight measured 0.14 in both eyes. The parent / carer reported the tests were “easy” to complete. Dilated bio microscopy, cycloplegic refraction, and disc and macular OCT found no ocular abnormality nor refractive error for this participant.

## Discussion

In this study, we compare the accuracy of parent-led, tablet-based VA tests to clinical standard, and orthoptic-led, tablet-based tests. Parents were left unsupervised in a private room to simulate a home environment. They were shown how to use the iSight app, maintain the correct testing distance, but were not given any live feedback on their testing technique. This approach resulted in measurements that could not be compared to clinical measures. However, they were comparable to measures collected by professionals using the same equipment, suggesting that equipment as well as testing technique plays a role in this difference.

There have been a variety of teams working to develop new equipment and methods of testing children’s visual acuity in a home setting. The *Amblyopia Tracker App* (Kay Pictures) and DigiVis are apps that attempt to control the variables of a typical visual acuity test by only allowing users to alter distance and not optotype size, and by measuring distance with a second device respectively (8,9). Both have good agreeability with clinical standard tests but further work on their utility and implementation is required. The *Peekaboo Vision* and *OKKO health* are apps gamify the visual acuity test. In Peekaboo Vision, children are presented with a grey screen with a grating stimulus in one corner (10,11). When the child touches the stimulus, they are rewarded as the stimulus transforms through animation into a smiling face. The test is made progressively harder through finer gratings until visual acuity threshold. Children appear to enjoy this method and it may have utility in visual acuity tests for children with Special Educational Needs. It is currently unknown which methods may encourage families to complete home visual acuity testing (12).

Families appear reluctant to use the apps at home. Painter et al. (2021) (13) contacted 103 parents or carers by telephone, inviting them to use a VA test at home with their children prior to their outpatient appointment. 96 families confirmed they would take part, but only 15 families (14.6%) completed the test and returned the results. Common reasons for not completing were lack of time or did not understand / receive the written instructions by email. Thirunavukarasu et al. (2021) (7) had similar problems with their DigiVis app, inviting 511 patients to take part in a research study of home vision tests, with 120 responding and participating (23%). DigiVis requires two devices, which may exclude some families that do not have access to equipment.

Access to equipment could be an important barrier to implantation of home vision testing. Children from the lowest socio-economic class are 1.82x more likely to have amblyopia (1) and may be less likely to adhere to current therapies (14,15). When planning service provision, policymakers should target this group, which are the least likely to have access to expensive equipment. All respondents to our parent / carer questionnaire had access to at least one device capable of running a VA test app at home, suggesting it unlikely that access to equipment is a significant barrier to use of the tests. Furthermore, parents and carers appeared to appreciate the usefulness of home-collected data and did not feel the test process would be challenging to do at home. All our participants received one to one demonstration of the app immediately prior to using it themselves. It appears that one-to-one demonstration may be a facilitator to home use, while written information (delivered by email) is less effective. Future studies should look at offering parents / carers a demonstration in the clinic with the test completed later at home. Further qualitative work to evaluate the process of home VA tests may identify areas for improvement in the implementation of these tests.

Clinicians and services also have reservations about widespread use of home vision tests. In June 2020, The Royal College of Ophthalmologists and British and Irish Orthoptic Society (BIOS) published a joint statement warning their members: “The reliability of apps when used by a parent or guardian in the home setting to test visual acuity in children is not yet proven” (5). A lack of reliability could lead to patients receiving appointments unnecessarily or not being seen when they ought to have been. Our data does little to absolve this notion with differences between clinical standard and home iSight tests likely caused by a combination of limited parent/carer training, outliers, and differences in equipment between the home and clinic tests.

In summary, our data suggests that while some parents/carers can test their children’s visual acuity using a clinic-like app, some may struggle. Erroneous measures may not always be possible to detect and as such we do not think these tests should be used without supervision from a professional, and with limited training only. Newer generations of tests are emerging that can control variables inherent to traditional visual acuity tests and/or gamify the process. It remains to be seen which approach becomes favoured for implementation into clinical practice. We highlight the need for tests to engage patients so as to increase the rate of uptake particularly among families from lower socio-economic backgrounds. Additionally, researchers and developers should be mindful of what equipment is required for patients to access new visual acuity tests.

## Data Availability

All data produced in the present study are available upon reasonable request to the authors

## Conflicts

No author has any conflict of interest, financial or otherwise, to declare. DO is funded by a National Institute for Health Research Pre-doctoral Clinical Academic Fellowship and equipment for the study was purchased by Gift of Sight (Southampton, UK).

---

| **Question** | **Response** | **N (%)** |
| --- | --- | --- |
| **Do you think you could do the test at home with your child?** | Yes, N (%) | 40 (97.6) |
|  | No, N (%) | 1 (2.4) |
| **Which of the following devices do you have daily access to at home?**<br>(Select all that apply) | Apple iPad | 18 (23.4) |
|  | Android tablet | 20 (26.0) |
|  | Android smartphone | 14 (18.2) |
|  | Apple iPhone | 23 (29.9) |
|  | Windows Smartphone | 2 (2.6) |
| **How easy did you find doing the visual acuity test with your child?** | Very difficult | 0 (0.0) |
|  | Difficult | 1 (2.4) |
|  | Easy | 21 (51.2) |
|  | Very easy | 19 (46.3) |
| **How helpful would you find the information from a home visual acuity assessment through the course of your child's treatment?** | Very unhelpful | 0 (0.0) |
|  | Unhelpful | 2 (4.9) |
|  | Helpful | 24 (58.5) |
|  | Very helpful | 15 (36.5) |

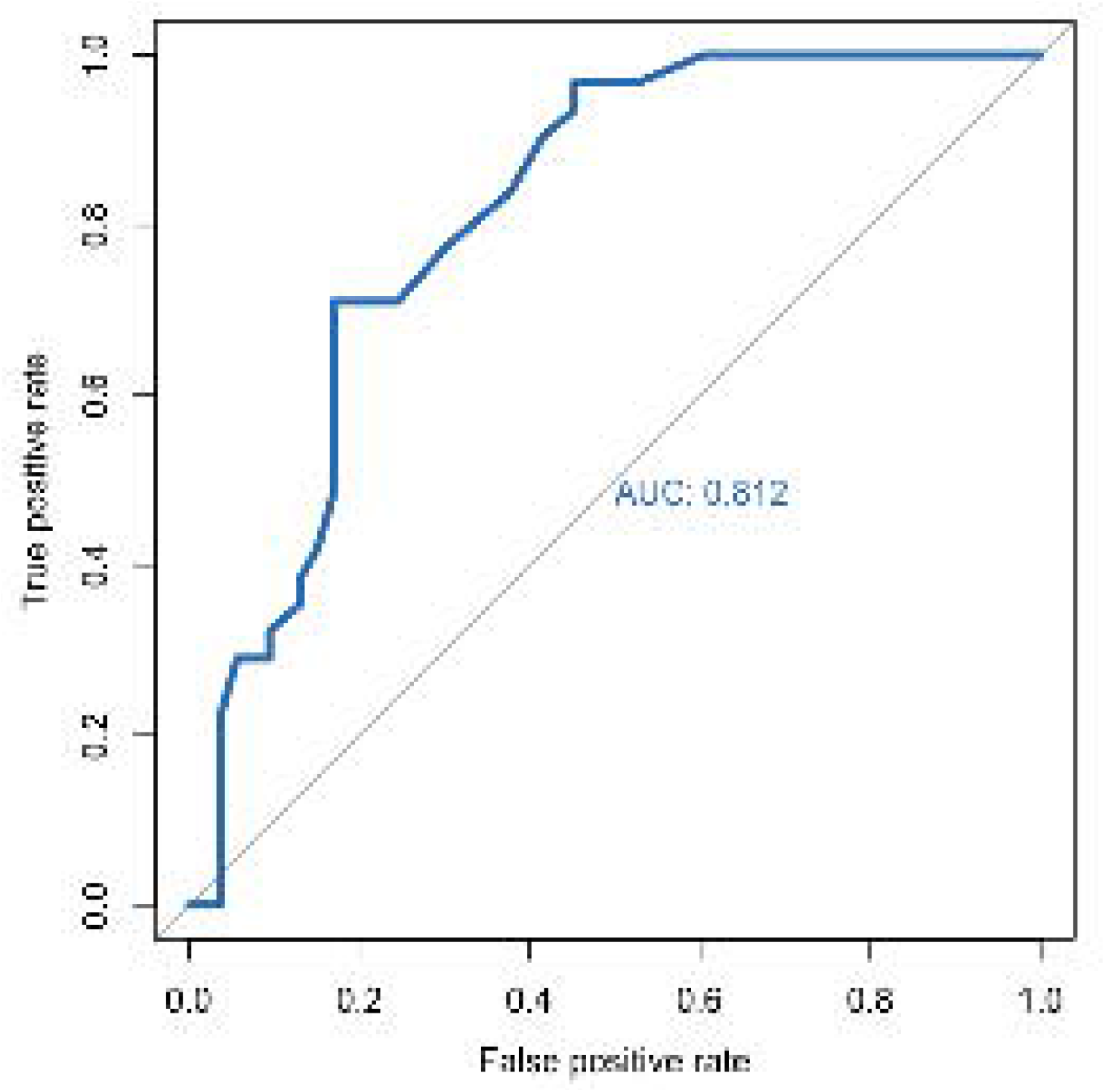

## Notes

### Competing Interest Statement

The authors have declared no competing interest.

